# Mapping the Mind in the Virtual Metaverse: An Initial In-Depth Thematic Exploration of Youth Mental Health within VRChat

**DOI:** 10.1101/2024.09.28.24314456

**Authors:** Tevin S Um, Raymond L Ownby, Shinnyi Chou

**Affiliations:** Department of Psychiatry, Nova Southeastern University Dr. Kiran C. Patel College of Osteopathic Medicine, Fort Lauderdale, FL, USA; Department of Psychiatry, University of Pittsburgh, Pittsburgh, PA, USA

## Abstract

**Background:** VRChat is a virtual reality (VR) social video game where users play a personalized avatar and socialize with others in community-built worlds. Its immersive nature and diverse community may impact youth users’ mental health, yet no research has explored this. This study is a thematic exploration of VRChat’s effect on youth mental health.

**Methods:** Semi-structured interviews were conducted with 20 active VRChat users aged 18-24 years. Interviews were transcribed and analyzed using grounded theory principles to identify themes related to VRChat use and mental health.

**Results:** Nine major themes emerged: Frequency of use; purpose of use; positive effects on mental health; negative effects on mental health; social connection and COVID-19; anonymity; cyberbullying and toxic behaviors; addiction; and recommendations for a safer environment. Positive effects identified included enhanced social connection, creative expression, and self-confidence. Negative effects included addiction, cyberbullying, predatory behaviors, and worsening of existing mental illnesses. The anonymity of VRChat created a paradoxical environment that can foster both genuine self-expression and toxic behaviors. During the COVID-19 pandemic, VRChat served as an important substitute for real-life social interactions. Participants also provided recommendations for developers, parents, and clinicians to promote a safer VRChat experience.

**Conclusion:** The immersive and interactive social experience offered by VRChat is conducive for positive effects on youth users’ mental health, such as social connectivity and self-expression. However, it also presents risks such as addiction and toxic behaviors. Given the interactive nature of VRChat, further research may elucidate its potential as a psychotherapeutic intervention for youth mental health.

## Introduction

Youth suffering from mental health issues such as depression and anxiety are at greater risk of experiencing academic failure, behavioral problems, and social deficits.^1^ In recent years, increased attention has been paid to how social media and online gaming impact youth mental health, and how digital platforms may be used to address mental health needs. ^2,3^ VRChat is an immersive and interactive game where users play as a personalized avatar and can join a room with other users to socialize. Interestingly, the immersive nature and accessibility of the platform make VRChat a potential intervention tool for youth mental health. However, no research has been done on VRChat and its effects on youth mental health, and gaining a better understanding of this topic may help guide both prevention and therapeutic intervention development. Thus, the present qualitative study aimed to provide a preliminary understanding of the impacts of VRChat on youth mental health. We hypothesized that VRChat usage has both positive and negative effects on youth mental health and that youth are interested in engaging with other stakeholders to improve the safety of VRChat usage, maximize its beneficial effects on mental health, and minimize potential harm.

## Materials and Methods

### Subjects

Participants from various demographic backgrounds were recruited between July and September 2023 to achieve population diversity. VRChat users were contacted randomly within VRChat’s preexisting “rooms” (instances of a pre-created map that users can create and join). After a user agreed to learn more about the study following a brief introduction, consent information was given, and all questions were answered before users consented to participate. A single male researcher (first author) conducted all interviews, and interviewer identity was anonymized through the standard default VRChat avatar and randomized username. All procedures were approved by the Institutional Review Board at Nova Southeastern University.

Inclusion criteria were (1) active VRChat users who had been VRChat users for at least three months and (2) participant age between 18 and 24 years. This age range was chosen because of the higher likelihood of experiencing mental health disorders such as anxiety and depression.^4^ Exclusion criteria included participants who could not engage appropriately to provide meaningful data analysis content. Individuals who self-identified as with or without a mental health disorder were recruited.

### Data Collection

Data was collected via a 30-minute semi-structured interview (see supplemental methods for interview script). Data saturation with the participants was discussed once no new recurring themes were discovered. To safeguard the participants’ anonymity, interviews with only the primary researcher and participant present were conducted through Zoom (Zoom Video Communications, Inc., Denver, U.S.A.) without video capture, and only audio recordings were collected via OBS Studio (Lain Bailey, https://obsproject.com/). Interviews were transcribed via Zoom’s built-in transcription feature, and audio files were destroyed immediately after transcribing. Transcripts for comment and/or correction were available for participants to review upon request.

### Data Analysis

Per the principles of Grounded Theory, the interview transcripts were input into QDA Miner Lite software (Provalis Research, Montreal, Quebec, Canada) for data analysis by the first author.^5^ Codes for each theme and subtheme of the interview were assigned within the software by a single coder (codes available upon request). Code frequency, participant checking, and constant comparative analysis of the contents were conducted. The guidelines for COnsolidated criteria for REporting Qualitative research (COREQ) were followed throughout the study (see supplemental methods for COREQ checklist).^6^

## Results

### Participant demographic

A total of 20 active VRChat users who met the inclusion criteria participated in this study (table 1). Five individuals declined to participate, citing lack of interest as the main reason. The mean age of participants was 21.5 ± 2.29 years (19.21 – 23.79). Participants’ gender self-identifications included 9 males (45%), 6 females (30%), and 5 non-binary/other individuals (25%). Fourteen (70%) participants had no known mental health diagnosis, and 6 (30%) participants had been diagnosed with one or more mental illnesses.

### Major themes and interpretations

Nine major themes were identified after analysis regarding the effects of VRChat on youth mental health. Table 2 provides major themes, minor themes, and representative quotations. Each theme and subtheme exploration is followed by relevant discussion points.

#### Theme 1: Frequency of Use

##### Daily and/or near-daily

Daily users (n = 13) can be subdivided into two groups. The first group comprised VRChat users who had deeply integrated the game into their lives, often blurring the lines between reality and virtual reality (i.e., “*it’s like a second home…*”). The second group had a similar frequency of usage. However, these users clearly distinguished their lives in VRChat from their day-to-day activities, with boundaries between virtual and “real-life” friends.

##### Weekly and Biweekly or more

The “Weekly” user group (n = 5) comprised casual consumers using the platform for relaxation. For those in the “Biweekly or more” group (n = 2), VRChat use was seen as an occasional event rather than a daily activity. Both groups reported less attachment to their VRChat experiences. They were less likely to mention the platform having a significant impact on their mental health, whether positive or negative.

#### Theme 2: Purpose of Use

##### Socialization and Relaxation

All participants reported a sense of community and belonging in VRChat. The deep friendships and community made on the platform were seen as “genuine” from a real-life human interaction standpoint. VRChat friend groups can do many of the same things that “real-life” friends can do, such as explore new places (e.g., visiting different VRChat worlds), participate in activities (e.g., attending VRChat dance parties), or sit in a circle conversing about life. Participants also reported using VRChat to escape their daily stress. These individuals found the same mental break from the calming environments and easygoing social interactions in VRChat as reading a book or watching a movie.

##### Escaping Reality

Several participants (n = 13) reported that VRChat was a tool for temporarily tuning out real-life challenges and escaping reality. They mentioned the psychological relief they experienced by “becoming someone else” in VRChat. This was achieved by pretending to be completely different from themselves, such as assuming a different persona, using an avatar that looks nothing like themselves in real-life, and using a pseudonym.

#### Theme 3: Positive Effects on Mental Health

##### Social connection

Most participants (n = 17) expressed enhanced social lives attributed to VRChat by maintaining preexisting friendships made in real-life and forging new friendships. Many users stated that the relationships they formed within VRChat translated to emotional support for real-life challenges. A notable point was the strengthening of long-distance relationships. For example, some users reported that VRChat was the primary method of communication with their significant other due to the addition of an immersive 3-dimensional element, as opposed to a 2-dimensional video call, allowing for an experience closer to “hanging out” in real-life.

##### Self-Confidence and Self-Expression

Many participants (n = 13) noted that VRChat’s open-minded environment catalyzed personal growth, translating into real-life self-confidence. Additionally, some participants stated that VRChat offered an avenue to connect with similar-minded people who resided outside their locale (i.e., “*I found my people here [in VRChat] … the people that live where I live don’t like the same stuff I do…*”).

##### Creative Outlet

The main methods for which participants expressed their creativity were through designing avatars, creating worlds (e.g., the worlds used to create VRChat “rooms”), or collaborating with others on various projects (e.g., organizing parties and raves within VRChat). These individuals viewed VRChat as a medium for artistic expression and reported satisfaction from their creative endeavors.

#### Theme 4: Negative Effects on Mental Health

##### Depression and/or Anxiety

Some participants (n = 8) expressed that VRChat can trigger or amplify depression and anxiety. This subtheme was expressed by participants both with (n = 6) and without (n = 14) diagnosed mental illness. Importantly, some users with diagnosed mental illness (n = 6) stated that their symptoms worsened with VRChat use, especially when their motivation for usage was to escape reality.

##### Judgment and/or Disapproval

Many participants (n = 13) expressed feeling judged or disapproved by groups within VRChat they did not identify with, which negatively impacted their self-esteem. In particular, participants who identified with the group “weeb” (a slang term for a non-Asian – specifically a non-Japanese person – who is fascinated with Asian or Japanese culture) experienced significant feelings of judgment from other non-”weeb” individuals (n = 7).

##### Addiction

The concern of addiction, especially among participants who were daily and/or near-daily users, is explored in its own major theme below.

#### Theme 5: Social Connection and COVID-19

##### Enhanced Social Closeness

Many VRChat users (n = 16) found that the virtual environment replaced day-to-day social interactions and added a novel element of closeness during the COVID-19 pandemic. Virtual meetings, hangouts, and parties in VRChat existed before the pandemic, but their popularity increased significantly during quarantine because they helped alleviate isolation during social distancing measures.

##### Replacement of Physical Interaction

Several participants (n = 14) expressed that VRChat was a complete or near-complete replacement of physical contact during the pandemic. Although this did not necessarily mean users felt physical sensations, a small portion of participants (n = 2) reported feeling tactile sensations in VRChat. For example, one user said that when interacting with another avatar, “I can almost feel … their hair and skin.”

##### Reintegration Post-Pandemic

Some users (n = 11) viewed VRChat as a short-term solution and were eager to return to real-life social interactions, while others (n = 4) considered VRChat’s social interactions superior to real-life. Some participants (n = 5) also supplemented their virtual relationships with real-life meetups post-quarantine.

#### Theme 6: Anonymity

##### Ease of Forming Friendships

The veil of anonymity in VRChat enabled many participants (n = 17) to initiate friendships without preconceptions. Participants appreciated that the social interactions were often based on personality, which many reported as their strong suit.

##### Acceptance in VRChat Compared to Real-Life

This subtheme highlighted a paradox--while VRChat is a place where one could be anyone (or anything), many participants (n = 15) felt they could genuinely be themselves. The platform allowed users to assume any identity through avatars that might differ significantly (both physically and figuratively) from real-life, and this freedom to self-reinvent paradoxically created a space where users could genuinely express themselves. Participants reported authentically expressing their identities, interests, and emotions, including sharing personal secrets or engaging in conversations they would usually shy away from.

##### Avatars

Participants (n = 15) discussed how their choice of avatars was not only a medium of self-expression but also affected how others perceived them. Since avatars can be custom-made, they are seen as one of the primary methods of self-expression within VRChat.

#### Theme 7: Cyberbullying and Toxic Behavior

##### Cyberbullying

Although anonymity positively influenced social interactions and self-explorations for some participants, a portion of participants (n = 10) spoke about experiencing or witnessing cyberbullying. The anonymity offered by VRChat seemed to entice some users to engage in destructive and toxic behaviors they might otherwise avoid in real-life due to social and legal consequences.

##### Cliques and niches

Several participants (n = 12) noted adverse effects from certain cliques and niches. Although these exclusive groups can create a protective buffer against toxic behaviors for their members, they can sometimes be the source of the toxicity and can lead to collective bullying/harassment (e.g. “*It’s almost like an ‘us versus them’ … to exclude outsiders …”*).

##### Predatory behaviors

Sexual predatory behavior towards underaged users is uncommon but not unheard of in VRChat. Some participants (n = 3) reported witnessing predatory behaviors targeting underaged users (e.g., “… he kept bothering her and wouldn’t stop following her …”). These behaviors may be attributed to the lack of a robust age-screening method and disinhibition potentiated by anonymity within VRChat.

#### Theme 8: Addiction

##### Dependency and/or Addiction

A few participants (n = 5) described going through “genuine withdrawal symptoms” when unable to access the platform. Some stated that they felt difficulty regulating the time and energy spent on VRChat. A part of this group also expressed that VRChat directly or indirectly hurt their school, work, or other areas of their personal lives. Increasingly compulsive use patterns might be induced by the immersive nature of VRChat, which could serve as powerful hooks.

##### Impacts on Academics, Work, Self-care, and Real-life Responsibilities

Extensive VRChat use was linked to real-life neglect, and some participants (n = 6) admitted they had neglected their schoolwork, missed work deadlines, or ignored simple self-care.

#### Theme 9: Recommendations for a Safer Environment

Participants highlighted recommendations to developers, parents, and clinicians regarding VRChat and mental health, outlined in Table 3.

## Conclusion

This study explored VRChat’s impact on youth mental health. We found that social connectivity, creative expression, and self-acceptance provided significant mental health benefits for VRChat users. However, addiction, cyberbullying, and predatory behaviors within VRChat adversely affected youth mental health. The anonymity of VRChat fosters a paradoxical freedom to discover one’s genuine self. Anonymity, unfortunately, also allows cyberbullying and toxic behaviors to run unchecked. VRChat is a novel gaming and social media platform that requires more attention from developers, parents, and clinicians to maximize its potential positive effects on mental health while minimizing harm. Research on VRChat is severely limited, and additional work is required to explore its potential as a therapeutic intervention. Research has already demonstrated that VR-based treatments are effective for specific phobias, general anxiety disorder, social anxiety disorder, public-speaking anxiety, PTSD and treatment-resistant PTSD, autism spectrum disorder, ADHD, and schizophrenia.^7-15^ Given the platform’s unique social dynamics and immersive, socially interactive nature, VRChat provides potential as an untapped treatment modality, especially for social anxiety. Controlled studies with VRChat may offer valuable insights into its efficacy as a primary or supplemental therapy to traditional treatments.

## Supporting information

Semi-Structured Script

COREQ

## Data Availability

All data produced in the present study are available upon reasonable request to the authors

## Acknowledgments

The authors would like to extend our sincere thanks to all of the interviewees of this project, who graciously and vulnerably shared their stories.

## Authorship Confirmation/Contribution Statement (CRediT)

Tevin S Um: Conceptualization (lead); Methodology (lead); Formal analysis (lead); Investigation (lead); Writing – Original Draft (lead); Visualization (lead). Raymond L Ownby: Resources (supporting); Writing – Review & Editing (supporting); Supervision (supporting). Shinnyi Chou: Conceptualization (supporting); Resources (lead); Writing – Review & Editing (lead); Supervision (lead)

## References

1. Costello EJ, Foley DL, Angold A. 10-year research update review: the epidemiology of child and adolescent psychiatric disorders: II. Developmental epidemiology. J Am Acad Child Adolesc Psychiatry. 2006;45(1):8–25. doi:10.1097/01.chi.0000184929.41423.c0

2. Schou Andreassen C, Billieux J, Griffiths MD, et al. The relationship between addictive use of social media and video games and symptoms of psychiatric disorders: A large-scale cross-sectional study. Psychol Addict Behav J Soc Psychol Addict Behav. 2016;30(2):252–262. doi:10.1037/adb0000160

3. Teachman BA, Silverman AL, Werntz A. Digital Mental Health Services: Moving From Promise to Results. Cogn Behav Pract. 2022;29(1):97–104. doi:10.1016/j.cbpra.2021.06.014

4. Bitsko RH, Holbrook JR, Ghandour RM, et al. Epidemiology and Impact of Health Care Provider–Diagnosed Anxiety and Depression Among US Children. J Dev Behav Pediatr JDBP. 2018;39(5):395–403. doi:10.1097/DBP.0000000000000571

5. Chun Tie Y, Birks M, Francis K. Grounded theory research: A design framework for novice researchers. SAGE Open Med. 2019;7:2050312118822927. doi:10.1177/2050312118822927

6. Tong A, Sainsbury P, Craig J. Consolidated criteria for reporting qualitative research (COREQ): a 32-item checklist for interviews and focus groups. Int J Qual Health Care. 2007;19(6):349–357. doi:10.1093/intqhc/mzm042

7. Emmelkamp PMG, Meyerbröker K, Morina N. Virtual Reality Therapy in Social Anxiety Disorder. Curr Psychiatry Rep. 2020;22(7):32. doi:10.1007/s11920-020-01156-1

8. Wiederhold BK, Wiederhold MD. Virtual Reality Therapy for Anxiety Disorders: Advances in Evaluation and Treatment. American Psychological Association; 2005:viii, 225. doi:10.1037/10858-000

9. Klinger E, Bouchard S, Légeron P, et al. Virtual Reality Therapy Versus Cognitive Behavior Therapy for Social Phobia: A Preliminary Controlled Study. Cyberpsychol Behav. 2005;8(1):76–88. doi:10.1089/cpb.2005.8.76

10. Hinojo-Lucena FJ, Aznar-Díaz I, Cáceres-Reche MP, Trujillo-Torres JM, Romero-Rodríguez JM. Virtual Reality Treatment for Public Speaking Anxiety in Students. Advancements and Results in Personalized Medicine. J Pers Med. 2020;10(1):14. doi:10.3390/jpm10010014

11. Gonçalves R, Pedrozo AL, Coutinho ESF, Figueira I, Ventura P. Efficacy of Virtual Reality Exposure Therapy in the Treatment of PTSD: A Systematic Review. PLOS ONE. 2012;7(12):e48469. doi:10.1371/journal.pone.0048469

12. Kothgassner OD, Goreis A, Kafka JX, Van Eickels RL, Plener PL, Felnhofer A. Virtual reality exposure therapy for posttraumatic stress disorder (PTSD): a meta-analysis. Eur J Psychotraumatology. 2019;10(1):1654782. doi:10.1080/20008198.2019.1654782

13. Karami B, Koushki R, Arabgol F, Rahmani M, Vahabie AH. Effectiveness of Virtual Reality-based therapeutic interventions on individuals with autism spectrum disorder: A comprehensive meta-analysis. Published online February 12, 2020. doi:10.31234/osf.io/s2jvy

14. Bioulac S, Micoulaud-Franchi JA, Maire J, et al. Virtual Remediation Versus Methylphenidate to Improve Distractibility in Children With ADHD: A Controlled Randomized Clinical Trial Study. J Atten Disord. 2020;24(2):326–335. doi:10.1177/1087054718759751

15. du Sert OP, Potvin S, Lipp O, et al. Virtual reality therapy for refractory auditory verbal hallucinations in schizophrenia: A pilot clinical trial. Schizophr Res. 2018;197:176–181. doi:10.1016/j.schres.2018.02.031

