## Supplementary material for "Mapping the Mind in the Virtual Metaverse: An Initial In-Depth Thematic Exploration of Youth Mental Health within VRChat": Table 1

**Table 1: Demographic Information of Participants**

|  | *n* | *%* |
| --- | --- | --- |
| Age |  |  |
| 18-20 | 8 | 40 |
| 21-24 | 12 | 60 |
| Gender |  |  |
| Male | 9 | 45 |
| Female | 6 | 30 |
| Non-binary/Other | 5 | 25 |
| Has a mental health diagnosis |  |  |
| No | 14 | 70 |
| Yes | 6 | 30 |
| Depression | 5 | 25 |
| Anxiety | 4 | 20 |
| ADHD | 2 | 10 |
