## Supplementary material for "Mapping the Mind in the Virtual Metaverse: An Initial In-Depth Thematic Exploration of Youth Mental Health within VRChat": Table 2

**Table 2: Major Themes, Minor themes, and Illustrative Quotations**

| **Major themes** | **Minor themes** | **N (%)** | **Representative Quotations from Participants Without a Diagnosed Mental Illness** | **Representative Quotations from Participants With a Diagnosed Mental Illness** |
| --- | --- | --- | --- | --- |
| *Frequency of Usage* | Daily and/or near-daily | 13 (65) | “[...] I’m on VRChat almost every day, it’s like a second home [...]” *(male)* | “[...] Around eight hours a day on average [...]” *(male. Diagnosed depression)* |
|  | Weekly | 5 (25) | “[...] Usually on the weekends [...]” *(Non-binary/Other.)* | “[...] I’d say around every week whenever I have some free time [...]” *(female. Diagnosed anxiety and depression)* |
|  | Biweekly or more | 2 (10) | “[...] Not super often, mostly when I feel really bored or when my friends want me to get on [...]” *(male.)* | N/A |
| *Purpose of Usage* | Socialization | 20 (100) | “[...] there’s new and interesting people you meet sometimes, some of my funniest memories are with a random person I met here [...]” *(female.)* | “[...] I like to meet with my friends… they are my closest friends [...]” *(male. Diagnosed depression)* |
|  | Relaxation | 16 (80) | “[...] it’s my favorite way to relax and have some fun [...]” *(male.)* | “[...] it’s just a nice way to relax after a long day [...]” *(Non-binary/Other. Diagnosed anxiety and depression)* |
|  | Escaping reality | 13 (65) | “[...] it’s a break from real-life… [real-life] sucks sometimes [...]” *(male.)* | “[...] it’s like a different world [...]” *(male. Diagnosed depression)* |
| *Positive Effects on Mental Health* | Social connection | 17 (85) | “[...] I usually chill with my girlfriend on VRChat [...]” *(male.)* | “[...] Some of my best friends I’ve made on here and they’re lifelong friends [...]” *(female. Diagnosed anxiety and depression)* |
|  | Self-confidence and self-expression | 13 (65) | “[...] VRChat lets me be who I really am without real-life pressures and anxiety… it has definitely helped me accept who I am and kind of helps me express myself in the real world too [...]” *(male.)* | “[...] being around my friends [who share the same interests] let’s me be who I really am [...]” *(male. Diagnosed depression)* |
|  | Creative outlet | 3 (15) | “[...] I love creating avatars and worlds, it’s hard, but I spend tons of time doing it [...]” *(male.)* | N/A |
| *Negative Effects on Mental Health* | Depression and/or anxiety | 8 (40) | “[...] Sometimes I feel a little depressed… they’re there, but they’re not really there [...]” *(female.)* | “[...] I have bad days… [certain situations] make my anxiety worse [...]” *(Non-binary/Other. Diagnosed anxiety and depression)* |
|  | Judgment and/or disapproval | 13 (65) | “[...] I’m a weeb… but [non-weebs] make fun of me a lot… they don’t like my avatar or the fact that I cosplay or the way I talk [...]” *(female.)* | “[...] some people judge me and [my avatar], they think it’s weird that my avatar is a girl, but I am actually a guy [...]” *(Non-binary/Other. Diagnosed anxiety and depression)* |
|  | Addition | 5 (25) | “[...] I guess I’m a little addicted, but… it’s not like a drug or anything like that [...]” *(female.)* | “[...] I sleep too late sometimes [because of VRChat]… and I feel tired the next day [...]” *(male. Diagnosed depression)* |
|  | Social isolation | 7 (35) | “[...] it’s not like real-life friendships, it’s different… [real] friendships are more personable [...]” *(male.)* | “[...] all of my friends are in [VRChat], I don’t really have friends in real life [...]” *(Non-binary/Other. Diagnosed anxiety and depression)* |
| *Social connection during COVID-19* | Enhanced social closeness | 16 (80) | “[...] definitely during COVID… I was by myself… it helped me not go insane [...]”*(male.)* | “[...] my first party [in VRChat] was awesome… I still go to them all the time [...]” *(male. Diagnosed depression)* |
|  | Replacement of physical interaction | 14 (70) | “[...] I can almost feel [interacting with another avatar] … their hair and skin [...]”*(Non-binary/Other.)* | “[...] [VRChat] was a lifesaver [during the pandemic]… when we couldn’t meet [in real-life] [...]” *(male. Diagnosed depression)* |
|  | Reintegration post-pandemic | 11 (55) | “[...] I feel like if I [did not use] VRChat, It’d be really hard going back [...]” *(male.)* | “[...] I feel like VRChat is way better than something like FaceTime or calling [...]” *(Non-binary/Other. Diagnosed anxiety and depression)* |
| *Anonymity* | Ease of forming friendships | 17 (85) | “[...] No judgment just based on the way I look… it’s way easier making friends on here than it is in real life [...]” *(female.)* | “[...] it’s tough making friends for me in real life… making friends here is way easier [...]” *(male. Diagnosed depression)* |
|  | Acceptance in VRChat compared to real-life | 15 (75) | “[...] people can’t judge me on my physical appearance… it’s liberating [...]” *(male.)* | “[...] I found out who I am here [in VRChat]… even though my avatar and my name is nothing like me [in real-life] [...]” *(male. Diagnosed depression)* |
|  | Use of avatars | 15 (75) | “[...] I love going into avatar world and finding one that truly resonates with me [...]” *(Nonbinary/Other.)* | “[...] [My avatar] affects the way people see me… for good or bad [...]” *(Non-binary/Other. Diagnosed anxiety and depression)* |
| *Cyberbullying and toxic behaviors* | Experience and/or witness cyberbullying | 10 (50) | “[...] I’ve definitely seen bullying on [VRChat]… these guys were laughing at [a singer] [...]” *(female.)* | “[...] [Cyberbullying] is pretty common… I see it basically everyday [...]” *(male. Diagnosed depression)* |
|  | Cliques and niches | 12 (60) | “[...] there’s different groups. “weebs,” “trolls,” and normal people [...]” *(male.)* | “[...] a lot of my friends are weebs… we don’t really talk to other people [...]” *(Non-binary/Other. Diagnosed anxiety and depression)* |
|  | Predatory behaviors | 3 (15) | “[...] One time I did see an older dude being extremely creepy towards a very obviously underaged girl, he kept bothering her and wouldn’t stop following her [...]” *(male.)* | “[...] some people here are gross… guys try to talk to me even when I tell them to go away [...]” *(female. Diagnosed anxiety and depression)* |
| *Addiction* | Dependency and/or addiction | 5 (25) | “[...] Maybe I do spend too much time on here, but it’s the best way I can spend time with my friends [...]” *(male.)* | “[...] I definitely do spend a lot of time on [VRChat]… I just feel like I need to [...]” *(male. Diagnosed depression)* |
|  | Impacts on academics, work, self-care, and real-life responsibilities | 6 (30) | “[...] when I play [VRChat] too late I’m really tired at work the next day [...]” *(Nonbinary/Other.)* | “[...] I have missed some [school] assignments because I’m on here so much, I just lose track [...]” *(Non-binary/Other. Diagnosed anxiety and depression)* |
| *Recommendations for a safer environment* | Suggestions for developers | 18 (90) | “[...] maybe a mental health resource could help [...]” *(female.)* | “[...] a better reporting system… so repeat offenders will think twice [...]” *(male. Diagnosed depression)* |
|  | Suggestions for parents | 12 (60) | “[...] Parents are just scared of what they don’t know. if they understood VRChat more, they could see that it’s not bad [...]” *(male.)* | “[...] Parents should watch their kids and make sure it’s safe [...]” *(male. Diagnosed depression)* |
|  | Suggestions for clinicians | 14 (70) | “[...] some kind of therapy, I feel like it’d be better than zoom calls. It’s more personable, not just a 2D video [...]” *(female.)* | “[...] it would be so cool to talk to my therapist in VRChat [...]” *(Non-binary/Other. Diagnosed anxiety and depression)* |
