## Supplementary material for "Mapping the Mind in the Virtual Metaverse: An Initial In-Depth Thematic Exploration of Youth Mental Health within VRChat": Table 3

Table 3: Recommendations to promote a safer VRChat environment provided by interview participants

| **Theme** | **Representative quotes** | **Recommendations summary** |
| --- | --- | --- |
| *For Developers* | | |
| Reporting and Monitoring | “[…] A better reporting system would be good… so repeat offenders will think twice about being toxic[…]” *(male. Diagnosed depression)* | A more robust reporting system would allow moderators to quickly and efficiently identify toxic and harmful behaviors. After identification of perpetrators of these behaviors, a warning system with subsequent chat restrictions or bans for repeated offenses would help diminish toxicity within the community. |
| Safe spaces | “[…] At the start of the game, people can be placed in like a safe space where they meet staff… this way people can learn more about the community and what’s acceptable or not […]” *(male.)* | An area where new users of VRChat can be welcomed by other new users and staff or volunteers can help players learn about the basics of the platform, including rules of what acceptable behaviors are. Opening these safe spaces, or creating separate safe spaces, to non-new users would allows any user to be in an environment that is more closely monitored and free from potential harassment. |
| *For Parents* | | |
| Education and Dialog | “[…] Parents need to know what their kids are getting into… all the good and the bad… it’s also their responsibility… they should educate themselves about VRChat […]” *(female.)* | Education for both the parents themselves and parents to children would allow for open dialog about VRChat. If parents are informed about the potential risks and benefits of a VR-based social platform, they will be able to convey the possible dangers of VRChat to their children while being vigilant. |
| *For Clinicians and Providers* | | |
| Technological Familiarity | “[…] Doctors should keep up with technology because it has such a big impact on everything … I would want a doctor that understands such a big part of my life […]” *(Non-binary/Other.)* | Immersive social media platforms like VRChat are rising in popularity, and clinicians should familiarize themselves with this growing sector. This will enable providers to understand the psychosocial dynamics of these platforms and allow more focused advice and therapy. |
| Screening | “[…] [VRChat] and other VR related questions should be asked about… [VR] is a big part of gaming and I know a lot of gaming communities can be toxic […]” *(male.)* | Questions related to online behaviors, experiences of cyberbullying or toxic behaviors, and peer-to-peer interactions within the virtual space can be included in routine assessments. With these screening questionnaires, clinicians can identify potential issues early. |
| Integrated Therapeutic Approaches | “[...] it would be so cool to talk to my therapist in VRChat [...]” *(Non-binary/Other. Diagnosed anxiety and depression)* | Virtual reality and VRChat could be incorporated into the clinical setting. Due to their immersive nature, they can be considered in therapies for social skills training, exposure therapy, and other interventions. However, potential risks and ethical considerations necessitate thorough evaluation first. |
| Multidisciplinary Teams | “[…] Developers, doctors, and therapists should be educated about VRChat, and about VR in general… everyone working together [in healthcare] has to be on the same page […]” *(male.)* | To tackle VRChat's challenges and potential holistically, professionals such as psychiatrists, technologists/developers, and ethicists should collaborate. Furthermore, research initiatives or developing guidelines for responsible usage may also be warranted. Given the unique challenges posed by virtual reality platforms such as VRChat, clinicians can advocate for ethical guidelines tailored to virtual interactions. These guidelines may inform both clinical practice and community guidelines. |
