## Supplementary material for "Mapping the Mind in the Virtual Metaverse: An Initial In-Depth Thematic Exploration of Youth Mental Health within VRChat": Semi-Structured Script

The Semi-Structured Script

*1. Can you tell me about your experience with VRChat? How often do you use it, and for what purposes?*

*2. In your opinion, what are the potential positive effects of VRChat on youth mental health? Have you experienced any of these effects personally?*

*1a. For example, how does VRChat make you feel, maybe feeling happy or excited?*

*2a. Do you feel closer to the people you talk to in VRChat?*

*3a. Do you feel the people you talk to accept and understand you?*

*3. On the other hand, what are the potential negative effects of VRChat on youth mental health? Have you experienced any of these effects personally or witnessed them in others?*

*1b. For example, how does VRChat make you feel, maybe feeling depressed or anxious? In what ways?*

*2b. Do you feel distant from the people you talk to in VRChat?*

*3b. Do you feel the people you talk to disapprove or judge you?*

*4. How do you think VRChat affects people’s emotions? Does it provide a safe space for expression and self-discovery, or does it make things like anxiety, depression, or loneliness worse?*

*5. Does VRChat help you feel socially closer to other people? Do you think it can help make people feel closer together, especially now after the COVID-19 pandemic?*

*6. How does not knowing who other people are and them not knowing who you are in VRChat affect acceptance and social interactions? For example, do you think forming friendships and finding acceptance in VRChat is easier than in real life?*

*7. Have you ever experienced or witnessed cyberbullying in VRChat? Cyberbullying is saying mean things that make other people feel bad. How do you think it affects you or other young people?*

*1c. If you were bullied, how did it make you feel?*

*2c. How did it change what you do?*

*8. Did you ever feel that you needed to be in VRChat, and couldn't stop yourself?*

*1d. Did you not do other important things, like school or taking care of yourself, to be on VRChat?*

*9. How does VRChat affect preexisting mental health disorders like anxiety, depression, or PTSD? Does it provide a coping mechanism or aggravate symptoms?*

*10. In your opinion, what steps can be taken to ensure that VRChat remains a safe and positive space for youth mental health? What can individuals, parents, and developers do to minimize the negative effects of VRChat on mental health?*
